# Supporting decision making for individuals living with dementia and their care partners with knowledge translation: an umbrella review

**DOI:** 10.1101/2024.09.17.24312581

**Authors:** Marie Biard, Flavie E. Detcheverry, William Betzner, Sara Becker, Karl S. Grewal, Sandi Azab, Patrick F. Bloniasz, Erin L. Mazerolle, Jolene Phelps, Eric E. Smith, AmanPreet Badhwar

**Author notes:** Corresponding author: AmanPreet Badhwar, MSc, PhD ^2^Multiomics Investigation of Neurodegenerative Diseases (MIND) lab, 4545 Chemin Queen May, Montréal, QC, H3W 1W4, Canada; ^3^Centre de Recherche de l’Institut Universitaire de Gériatrie de Montréal (CRIUGM), 4545 Chemin Queen May, Montréal, QC, H3W 1W4, Canada; ^4^Department of Pharmacology and Physiology, Université de Montréal, 2900 Boulevard Édouard-Montpetit, Montréal, QC, H3T 1J4, Canada; ^5^Institute of Biomedical Engineering, Université de Montréal, 2960 Chemin de la Tour, Montréal, QC, H3T 1J4, Canada., Postal address: 4565, chemin Queen Mary, Montreal, QC, Canada, H3W 1W5. indicates equal first co-authorship (Marie Biard, Flavie E. Detcheverry).

## Abstract

Living with dementia requires decision making about numerous topics including daily activities and advance care planning (ACP). Both individuals living with dementia and care partners require informed support for decision making. We conducted an umbrella review to assess knowledge translation (KT) interventions supporting decision making for individuals living with dementia and their informal care partners. Four databases were searched using 50 different search-terms, identifying 22 reviews presenting 32 KT interventions. The most common KT decision topic was ACP (N=21) which includes advanced care directives, feeding options, and placement in long-term care. The majority of KT interventions targeted care partners only (N=16), or both care partners and individuals living with dementia (N=13), with fewer interventions (N=3) targeting individuals living with dementia. Overall, our umbrella review offers insights into the beneficial impacts of KT interventions, such as increased knowledge and confidence, and decreased decisional conflicts.

## 1. INTRODUCTION

Dementia is an umbrella term including a wide range of specific medical conditions caused by abnormal brain changes. Alzheimer’s disease (AD) and vascular dementia are the two most prevalent forms of dementia, and sometimes co-exist as mixed dementia [1,2]. Over 55 million people worldwide currently live with dementia, a number projected to reach 139 million by 2050 [3]. While the presenting symptoms of dementia vary, most individuals experience mild cognitive impairment (MCI), years before the dementia diagnosis [4]. Eventually, the progressive cognitive decline associated with dementia impedes the individual’s autonomy and self-efficacy, generally requiring the involvement of family members or other informal care partners [3].

Assistance provided by informal care partners is estimated at around five hours per day per person with dementia [5], which can negatively impact care partners’ well being [6,7]. Since informal caregiving accounts for approximately half of the overall costs of dementia (total of 651.4 billion US$ in 2019 [8]), the World Health Organization addressed the need for support to care partners of individuals living with dementia in the global action plan on the public health response to dementia [6]. While some progress has been made since its approval in 2017, more is needed to support informal care partners [3].

Dementia care involves making decisions regarding several topics, from day-to-day activities (e.g., driving cessation, fall prevention, medication management) to end-of-life care [9,10]. While some individuals living with dementia prefer to rely on their family for decision making, others wish to participate in making their own decisions for as long as possible [11]. One way to support decision making of individuals living with dementia and their care partners is through the development of Knowledge Translation (KT) interventions.

KT is the process of creating, synthesizing, and applying knowledge with the aim of improving health and strengthening the healthcare system [12]. Examples of KT interventions for end-users, such as patients and care partners range from structured discussions with a healthcare professional to more standardized tools such as decision aids (e.g., booklets, pamphlets, audiovisual interventions), which have been shown to help patients make decisions, on topics such as treatment or screening decisions [13,14].

We conducted an umbrella review to assess existing KT interventions aimed at helping individuals living with dementia and care partners make decisions about current or future matters. Umbrella reviews are a relatively recent concept in evidence synthesis, addressing gaps in individual systematic reviews by providing a broad, up to date overview of a topic [15]. We specifically focused on KT interventions targeting the two most common forms of dementia, namely AD, vascular cognitive impairment and dementia (VCID; both the prodromal and dementia stages of vascular dementia), and mixed dementia. Our specific aims were to investigate: (a) What types of KT interventions exist for individuals living with dementia and their care partners to improve informed decision making? (b) What are the outcomes of existing KT decision support interventions for individuals living with dementia and their care partners? and (c) What gaps exist in KT about decision-making support for individuals living with dementia and their care partners?

## 2. METHODS

This umbrella review was conducted based on the Joanna Briggs Institute (JBI) methodology for umbrella reviews [16,17]. Prior to redaction, the study protocol was registered on PROSPERO (CRD42023414419; Supplementary Material S1).

### 2.1 Search strategy

Following a pilot search on MEDLINE, a search strategy addressing key concepts of our review question, namely KT, decision making, knowledge users, and dementia, was developed (M.B., E.L.M., E.E.S., A.B.). The full search strategy is available in the Supplementary Material S2. The search was conducted on January 7, 2023, using 50 different search terms in four electronic databases on Ovid: MEDLINE, APA PsycINFO, Embase, and Cochrane database of systematic reviews. No time period restriction was applied. Only English-written articles were considered, justified by findings from the pilot search that expanding to additional languages was unlikely to yield substantially more articles. The search was supplemented by citation tracking the references of the included studies as well as searching for grey literature such as reports from the Canadian Institute for Health Information. If review protocols or conference abstracts were retrieved, a search was carried out to check whether the findings were subsequently published as full length, peer-reviewed articles.

### 2.2 Inclusion and exclusion criteria

#### 2.2.1 Population

Literature syntheses focusing on individuals living with dementia or their informal care partners were included. Individuals living with any of the following major dementia subtypes were accepted: AD, VCID, and mixed dementia. Note that when provided, ethnicity information of the target population was extracted and specified. Reviews solely targeting individuals living with other dementia subtypes (e.g., Lewy Body dementia, Parkinson’s disease related dementia), individuals with other illnesses (e.g., stroke), or older adults in general were excluded. We further excluded studies focusing only on healthcare professionals (e.g., nurses), but included those with interventions directed at dyads (individual with dementia and informal care partner) or triads (individual with dementia, informal care partner, and health professional).

#### 2.2.2 Intervention

Systematic reviews synthesizing interventions that support decision making for individuals living with dementia and/or their care partners were included. For example, these could take the form of booklets, pamphlets, audiovisual interventions, structured discussion, or educational sessions.

#### 2.2.3 Comparator

No review article was excluded based on the comparator used (i.e., usual care), or the lack of comparator, given the broad nature of our umbrella review question and the *a priori* inclusion of both qualitative and quantitative studies.

#### 2.2.4 Outcome

Likewise, systematic reviews reporting any outcomes (e.g., increase in knowledge about dementia/dementia care and decision-making skills, satisfaction of quality of care, confidence) were included.

#### 2.2.5 Type of studies

We included review articles that used (a) a defined, reproducible search strategy, (b) systematically applied inclusion and exclusion criteria, and (c) produced a synthesis of findings, which may be qualitative. These reviews are usually self identified as: “Systematic review”, “Meta-analysis”, “Rapid review”, “Umbrella review”, and “Scoping review”. Pre-print reviews, conference abstracts, or review protocols were excluded.

### 2.3 Study screening and selection

The screening process was performed using the Covidence platform. Nine independent reviewers (M.B., F.E.D., S.A., S.B., W.B., P.F.B., K.S.G., J.P., and E.E.S.) participating in the Vascular Training (VAST) Platform collaborative initiative screened titles and abstracts against inclusion/exclusion criteria listed above. Every record was screened independently by M.B. and a second reviewer from the list of independent reviewers. Full-text review of included publications was independently examined by two reviewers (M.B. and E.E.S.), and conflicts were resolved by discussion, with the help of a third reviewer when necessary.

### 2.4 Critical appraisal

Two reviewers (M.B. and F.E.D.) independently conducted the critical appraisal of the selected systematic reviews using the JBI Critical Appraisal Checklist for Systematic Reviews and Research Syntheses [16,17]. The checklist included eleven questions, of which nine assessed the validity of the study. The last two questions assessed the relevance of the recommendations and gaps identified. Reviewers did a pilot critical appraisal to calibrate their responses at the beginning of the quality assessment.

### 2.5 Data extraction

Two independent reviewers (M.B. and F.E.D.) used a modified version of the JBI Data Extraction Tool [16,17]. Specifically, the following elements were extracted in a standardized manner from included reviews (review-level): title, author, year, country, type of review, population, comparator, intervention, outcomes, databases searched, appraisal instrument and rating, results, and other comments. While extracting data from reviews, relevant primary articles about interventions meeting our inclusion criteria (i.e., targeting individuals living with AD, VCID, or mixed dementia, and/or their care partners) were identified, resulting in 57 relevant primary articles. To ensure no information was missed about these KT interventions, we extracted the following information from the primary articles (individual study-level): title, author, year, country, type of review, intervention, outcomes, and appraisal instrument and rating. Reviewers completed a pilot data extraction beforehand to calibrate responses. Conflicts were resolved following the same process as outlined above.

### 2.6 Data summary

Given that the knowledge syntheses found in the literature were likely to be heterogeneous in terms of design and objectives, a narrative format was adopted to synthesize the results of this umbrella review.

## 3. RESULTS

### 3.1 Study inclusion

The search yielded a total of 2,265 studies after duplicate removal, of which 82 remained after title and abstract screening. After full-text assessment, 60 additional studies were excluded, resulting in 22 included reviews. Our Preferred Reporting Items for Systematic Reviews and Meta-Analyses (PRISMA) flow diagram is provided in Figure 1.

**FIGURE 1.**
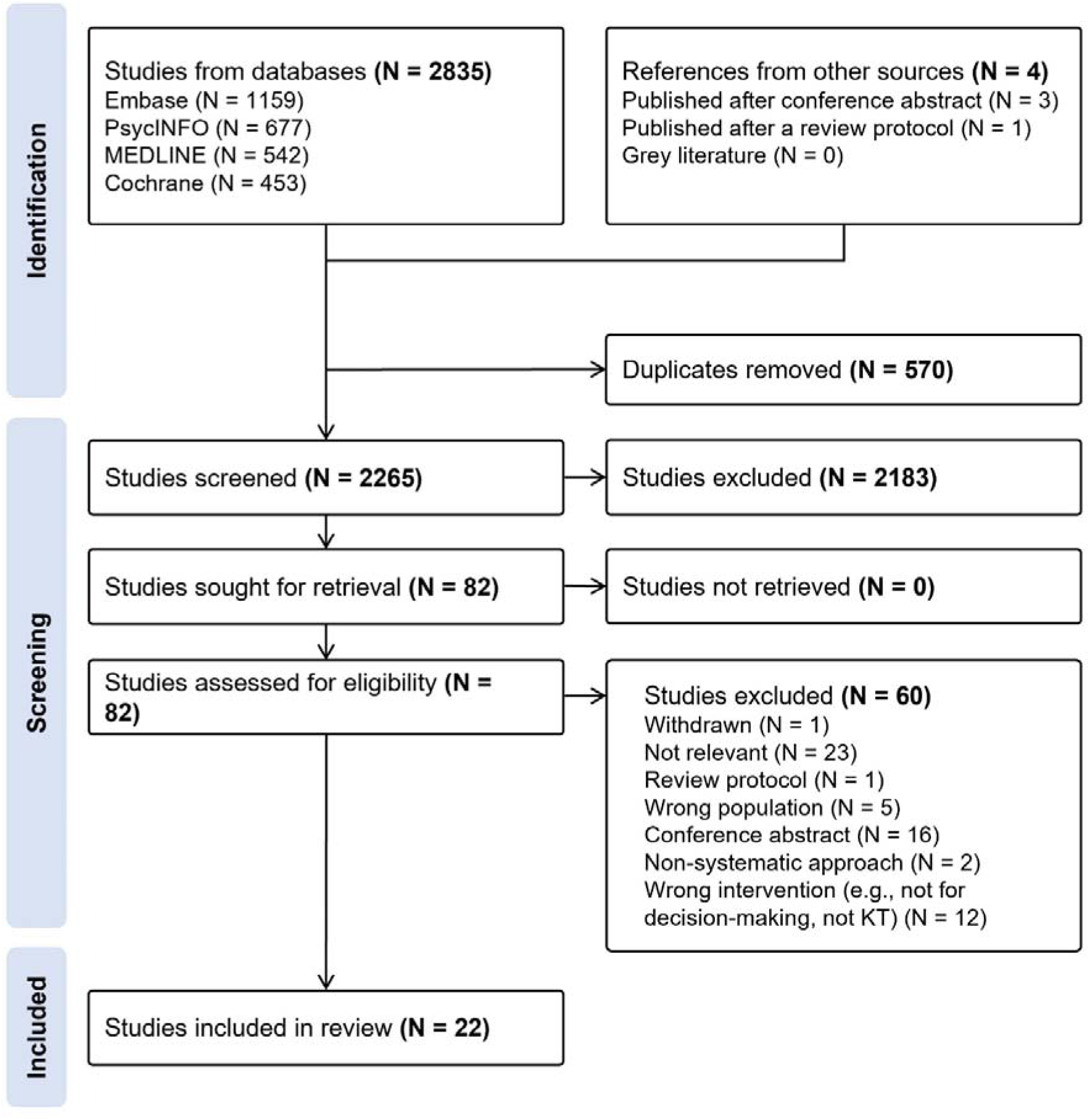
Preferred Reporting Items for Systematic Reviews and Meta-Analyses (PRISMA) flow diagram of the umbrella review selection process.

### 3.2 Methodological quality

The assessment of the risk of bias of our included studies was performed using the JBI Critical Appraisal Checklist for Systematic Reviews and Research Syntheses [16,17], and is displayed in Figure 2. According to the percentage of criteria met using the JBI critical appraisal checklist, 17 studies were of high quality, four were of medium quality, and one was of low qualities. For 9 of the 11 risk of bias questions (questions 1-5, 7, 8, 10, and 11), fewer than 25% of studies were rated as medium or high risk of bias. For question 6, which asked about whether critical appraisal conducted by two or more reviewers independently, more than 50% of studies were rated as medium to high risk of bias. For question 9, which asked about likelihood of publication bias, more than 50% of studies were rated as high risk of bias. Overall, the majority of studies clearly stated their review question, had appropriate inclusion criteria and search strategy, and used adequate methods for data extraction and synthesis.

**FIGURE 2.**
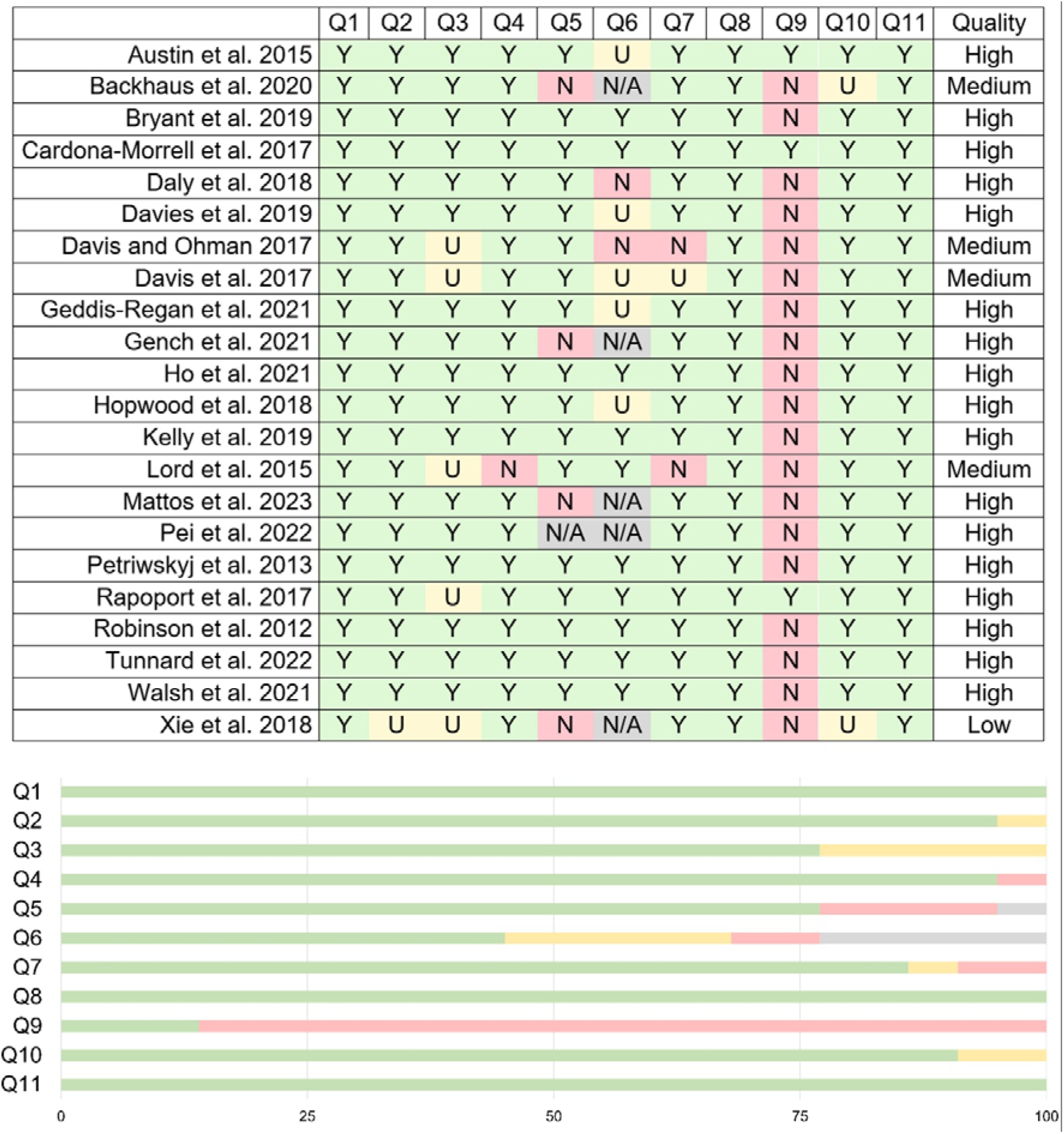
Critical appraisal for included reviews. % of criteria met: <50% high risk of bias; 50-75% medium risk of bias; >75% low risk of bias. *Abbreviations*: Y, Yes. N, No. U, Unclear. N/A, not applicable. Q1. Is the review question clearly and explicitly stated?; Q2. Were the inclusion criteria appropriate for the review question?; Q3. Was the search strategy appropriate?; Q4. Were the sources and resources used to search for studies adequate?; Q5. Were the criteria for appraising studies appropriate?; Q6. Was critical appraisal conducted by two or more reviewers independently?; Q7. Were there methods to minimize errors in data extraction?; Q8. Were the methods used to combine studies appropriate?; Q9. Was the likelihood of publication bias assessed?; Q10. Were recommendations for policy and/or practice supported by the reported data?; Q11. Were the specific directives for new research appropriate?.

### 3.3 Characteristics of included reviews

General study characteristics are presented in Table 1. There were a total of 22 reviews from 2012 to 2023 (Figure 3A), targeting individuals living with dementia, care partners, and healthcare professionals (Figure 3B), and including four KT intervention categories (Figure 3C). These studies were stemming from the United Kingdom (N=7) [18–24], Australia (N=6) [25–30], the United States of America (N=6) [31–36], Canada (N=1) [37], Ireland (N=1) [38], and the Netherlands (N=1) [39] (Figure 3D). Out of the 22 reviews identified, 18 were systematic reviews [18–28,30,31,35,37–40], two were scoping reviews [34,36], and two were integrative reviews [32,33] (Table 1). All used a narrative synthesis to report the results. Finally, two reviews additionally performed a meta-analysis [24,35].

**FIGURE 3.**
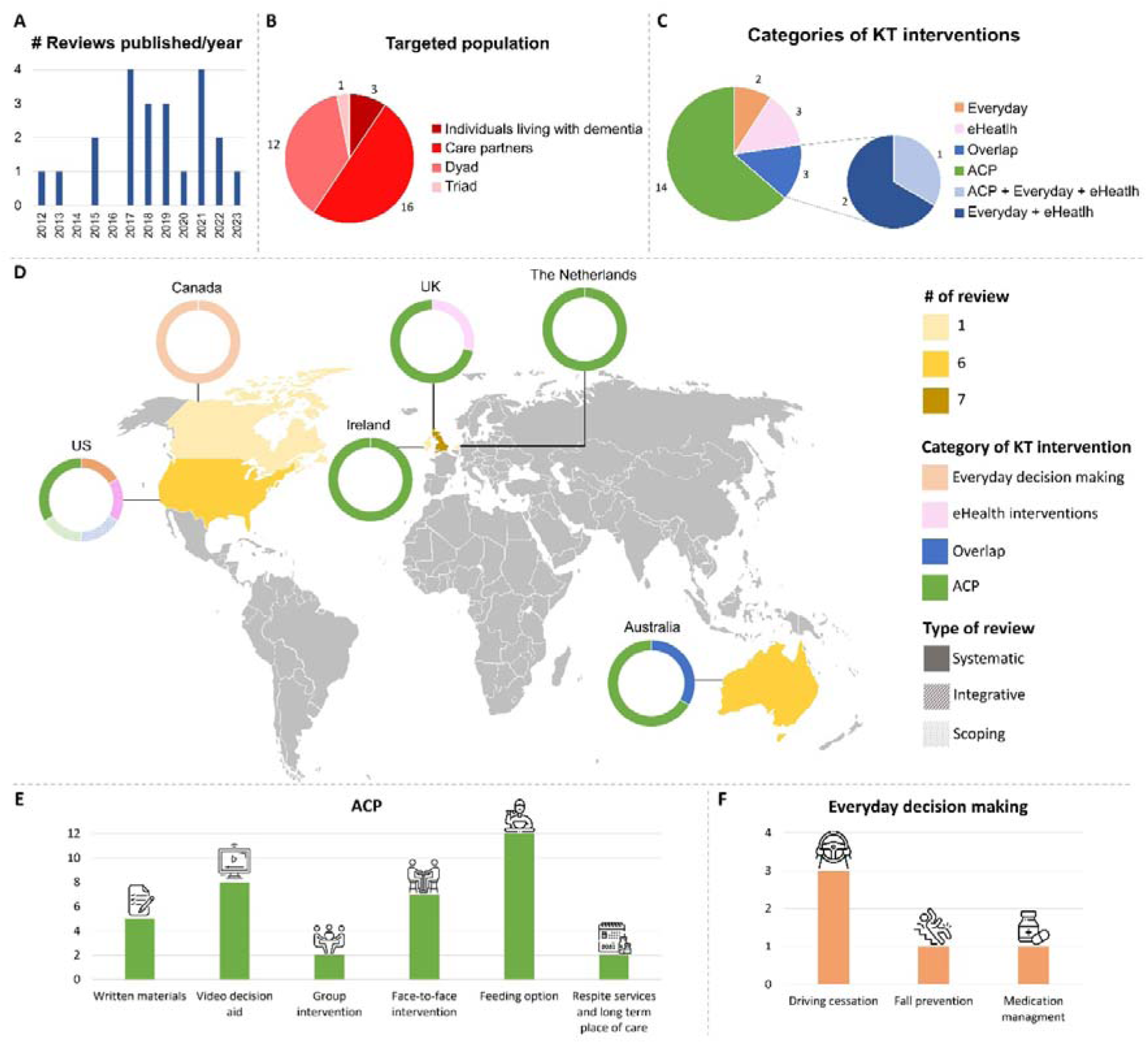
Description of included reviews and KT interventions. **A)** Publication years of included reviews, **B)** Targeted population of KT interventions, **C)** Categories of KT interventions, **D)** Geographical distribution of included reviews, **E)** Sub-categories of everyday decision-making interventions, and **F)** Sub-categories of advance care planning interventions. *Note:* Numbers provided for categories of KT interventions per included reviews, while numbers for targeted population are per interventions. *Abbreviations:* ACP, advance care planning; KT, knowledge translation.

**TABLE 1.** Characteristics of included reviews. *Abbreviations*: UK, United Kingdom; US, United States.

| Reference | Country | Type of review | Intervention | Population | Proportion of relevant primary studies |
| --- | --- | --- | --- | --- | --- |
| Austin et al., 2015 | US | Systematic | Support interventions for current or future care decisions | Mixed population: adults with advanced or terminal illness | 9/38 |
| Backhaus et al., 2020 | The Netherlands | Systematic | Interventions to facilitate the inclusion of family | Individuals living with dementia | 4/29 |
| Bryant et al., 2019 | Australia | Systematic | Intervention to improve advance care planning | Individuals living with dementia | 4/4 |
| Cardona-Morrell et al., 2017 | Australia | Systematic | Decision aids for end-of-life care | Mixed population: older adults with advanced or terminal illness | 3/17 |
| Daly et al., 2018 | UK | Systematic | Shared decision-making interventions | Mixed population: any form of cognitive impairment | 2/19 |
| Davies et al., 2019 | UK | Systematic | Decision aids | Individuals living with dementia | 10/10 |
| Davis and Ohman, 2017 | US | Integrative | Driving related interventions | Individuals living with dementia or mild cognitive impairment | 1/26 |
| Davis et al., 2017 | US | Integrative | Everyday decision-making related interventions | Individuals living with dementia | 1/12 |
| Geddis-Regan et al., 2021 | UK | Systematic | Support interventions for shared or surrogate care decisions | Individuals living with dementia | 6/8 |
| Gench et al., 2021 | Australia | Systematic | Medication management interventions | Individuals living with dementia | 2/10 |
| Ho et al., 2021 | Australia | Systematic | Decision-making tools | Individuals living with dementia or mild cognitive impairment | 14/35 |
| Hopwood et al., 2018 | UK | Systematic | Internet-based support interventions | Individuals living with dementia | 6/40 |
| Kelly et al., 2019 | Australia | Systematic | Advance care planning interventions | Individuals living with dementia | 16/30 |
| Lord et al., 2015 | UK | Systematic | Support interventions for surrogate decision-making | Individuals living with dementia | 3/30 |
| Mattos et al., 2023 | US | Scoping | Shared decision-making interventions | Individuals living with dementia | 6/32 |
| Pei et al., 2022 | US | Scoping | Decision aids about feeding options | Individuals living with dementia | 6/6 |
| Petriwskyj et al., 2013 | Australia | Systematic | Family involvement in decision-making | Individuals living with dementia | 2/26 |
| Rapoport et al., 2017 | Canada | Systematic | Support interventions for driving cessation | Mixed population: older adults with or without dementia | 1/3 |
| Robinson et al., 2012 | UK | Systematic | Advance care planning interventions | Individuals living with dementia | 2/4 |
| Tunnard et al., 2022 | UK | Systematic | eHealth interventions for care decision-making | Individuals living with dementia | 2/26 |
| Walsh et al., 2021 | Ireland | Systematic | Palliative care interventions | Individuals living with dementia | 4/9 |
| Xie et al., 2018 | US | Systematic | Decision aids for end-of-life care | Individuals living with dementia | 5/5 |

While all the selected reviews included primary studies of individuals living with the conditions (i.e., AD, VCID, or mixed dementia) that were specified in our selection criteria, most of them (18 of 22) also included studies of patients with other conditions. This was due to the fact that the majority of reviews had included additional objectives not specifically involving decision-making interventions or had targeted a mixed population of individuals living with dementia or other conditions. Altogether, the 22 reviews reported a total of 57 unique primary articles relevant to our aims (Supplementary Material S3). We were unable to assess one primary article [41], which was not available in English.

### 3.4 Findings

Findings are reported based on three main KT intervention categories: (a) Advance Care Planning (ACP) (Section 3.4.1.), (b) everyday decision making (Section 3.4.2.), and (c) eHealth interventions (Section 3.4.3.). In addition, we have summarised findings relative to KT interventions targeted at care partners only (N=16), dyads (N=12) or triads (N=1), and individuals living with dementia only (N=3), in Tables 2, 3, and 4, respectively.

**TABLE 2.**
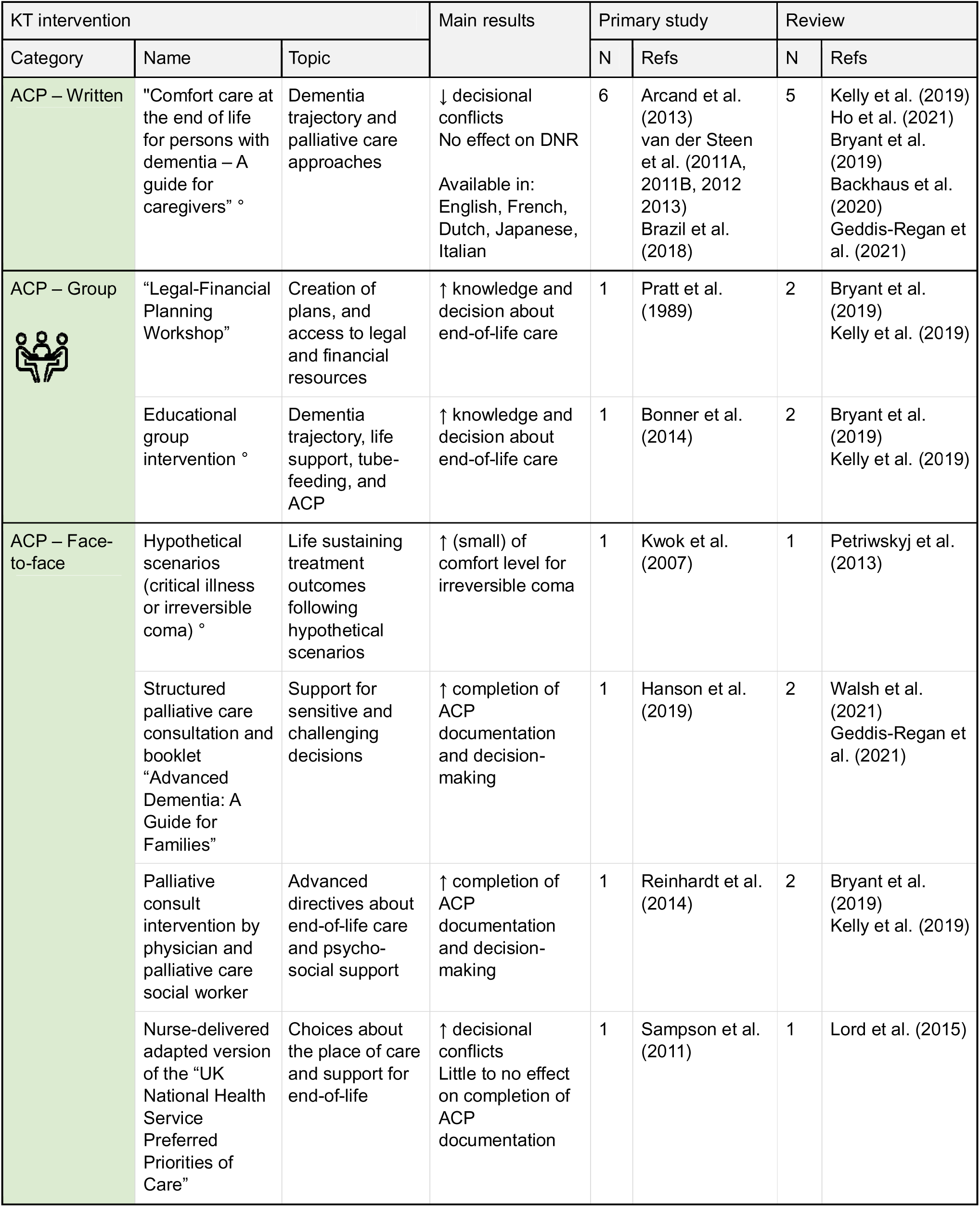
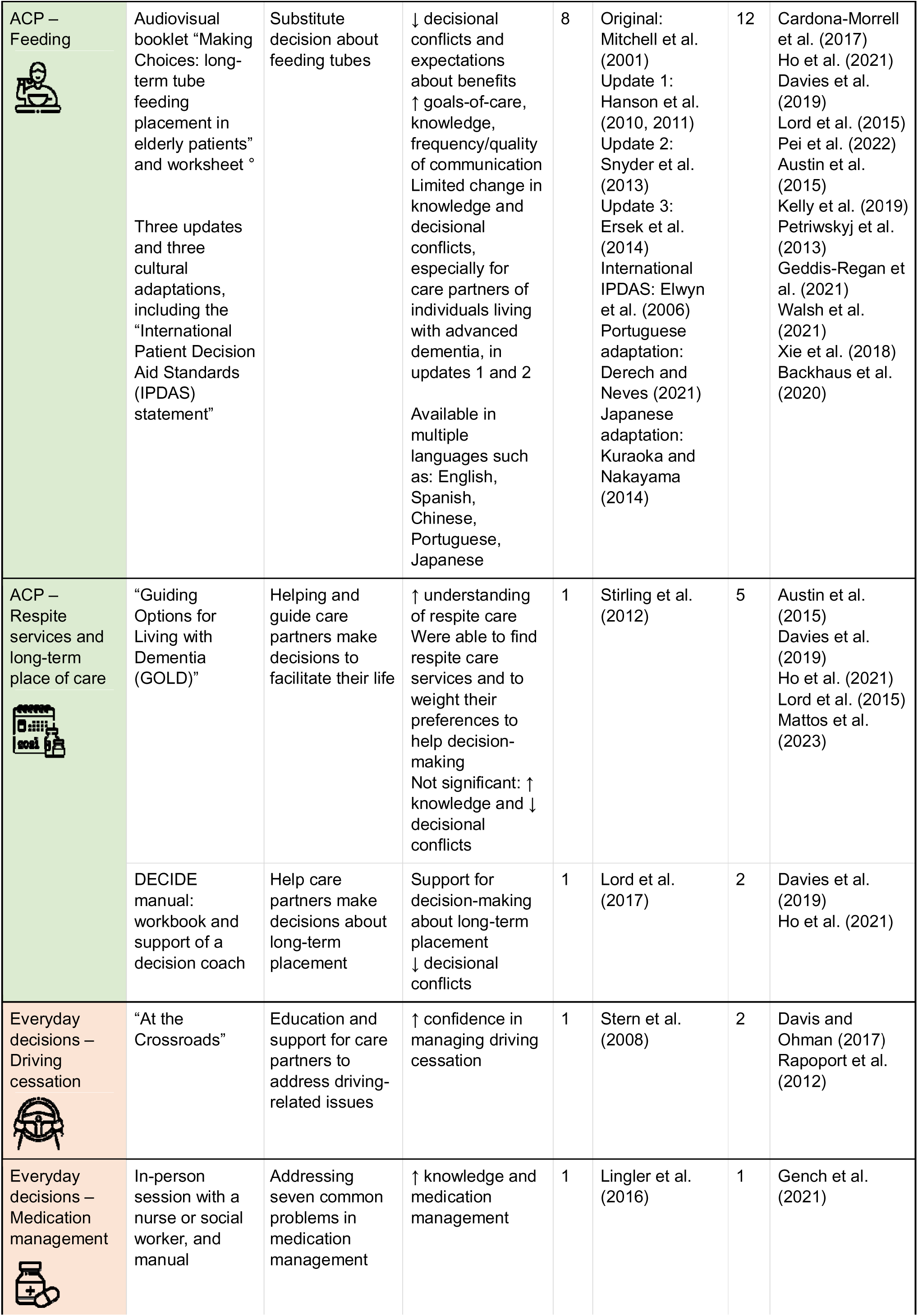
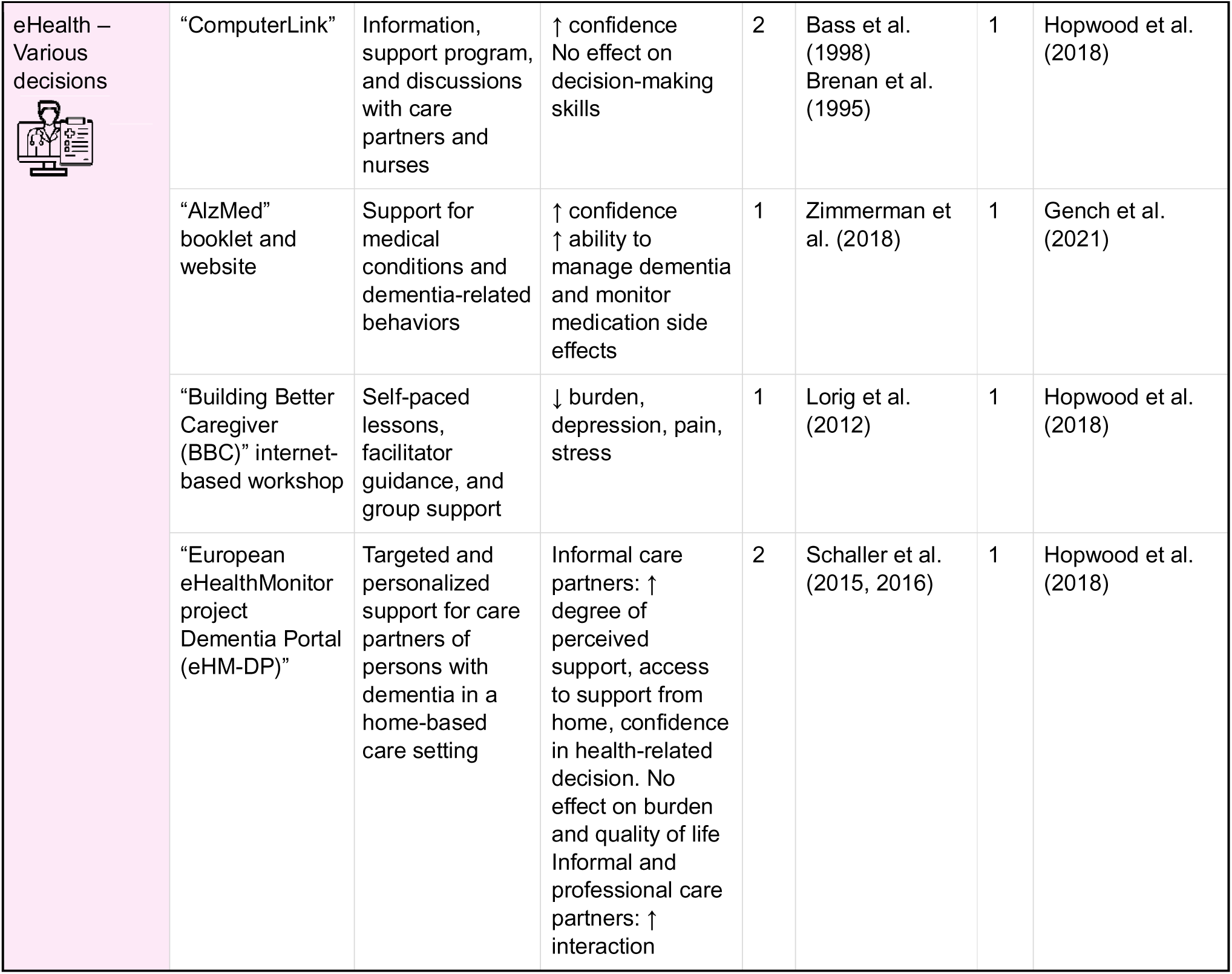
Interventions targeting care partners. Note that the original version of the audiovisual booklet *“Making Choices: long-term tube feeding placement in elderly patients”* and worksheet targeted older individuals without dementia. *Abbreviations*: ° multiple languages and/or cultural adaptations available; ACP, advance care planning; KT, knowledge translation.

| KT intervention |  |  | Main results | Primary study |  | Review |  |
| --- | --- | --- | --- | --- | --- | --- | --- |
| Category | Name | Topic |  | N | Refs | N | Refs |
| ACP – Written | "Comfort care at the end of life for persons with dementia – A guide for caregivers" ° | Dementia trajectory and palliative care approaches | ↓ decisional conflicts<br>No effect on DNR<br>Available in: English, French, Dutch, Japanese, Italian | 6 | Arcand et al. (2013)<br>van der Steen et al. (2011A, 2011B, 2012 2013)<br>Brazil et al. (2018) | 5 | Kelly et al. (2019)<br>Ho et al. (2021)<br>Bryant et al. (2019)<br>Backhaus et al. (2020)<br>Geddis-Regan et al. (2021) |
| ACP – Group | "Legal-Financial Planning Workshop" | Creation of plans, and access to legal and financial resources | ↑ knowledge and decision about end-of-life care | 1 | Pratt et al. (1989) | 2 | Bryant et al. (2019)<br>Kelly et al. (2019) |
|  | Educational group intervention ° | Dementia trajectory, life support, tube-feeding, and ACP | ↑ knowledge and decision about end-of-life care | 1 | Bonner et al. (2014) | 2 | Bryant et al. (2019)<br>Kelly et al. (2019) |
| ACP – Face-to-face | Hypothetical scenarios (critical illness or irreversible coma) ° | Life sustaining treatment outcomes following hypothetical scenarios | ↑ (small) of comfort level for irreversible coma | 1 | Kwok et al. (2007) | 1 | Petriwskyj et al. (2013) |
|  | Structured palliative care consultation and booklet "Advanced Dementia: A Guide for Families" | Support for sensitive and challenging decisions | ↑ completion of ACP documentation and decision-making | 1 | Hanson et al. (2019) | 2 | Walsh et al. (2021)<br>Geddis-Regan et al. (2021) |
|  | Palliative consult intervention by physician and palliative care social worker | Advanced directives about end-of-life care and psycho-social support | ↑ completion of ACP documentation and decision-making | 1 | Reinhardt et al. (2014) | 2 | Bryant et al. (2019)<br>Kelly et al. (2019) |
|  | Nurse-delivered adapted version of the "UK National Health Service Preferred Priorities of Care" | Choices about the place of care and support for end-of-life | ↑ decisional conflicts<br>Little to no effect on completion of ACP documentation | 1 | Sampson et al. (2011) | 1 | Lord et al. (2015) |
| ACP – Feeding | Audiovisual booklet “Making Choices: long-term tube feeding placement in elderly patients” and worksheet °<br>Three updates and three cultural adaptations, including the “International Patient Decision Aid Standards (IPDAS) statement” | Substitute decision about feeding tubes | ↓ decisional conflicts and expectations about benefits<br>↑ goals-of-care, knowledge, frequency/quality of communication<br>Limited change in knowledge and decisional conflicts, especially for care partners of individuals living with advanced dementia, in updates 1 and 2<br>Available in multiple languages such as: English, Spanish, Chinese, Portuguese, Japanese | 8 | Original: Mitchell et al. (2001)<br>Update 1: Hanson et al. (2010, 2011)<br>Update 2: Snyder et al. (2013)<br>Update 3: Ersek et al. (2014)<br>International IPDAS: Elwyn et al. (2006)<br>Portuguese adaptation: Derech and Neves (2021)<br>Japanese adaptation: Kuraoka and Nakayama (2014) | 12 | Cardona-Morrell et al. (2017)<br>Ho et al. (2021)<br>Davies et al. (2019)<br>Lord et al. (2015)<br>Pei et al. (2022)<br>Austin et al. (2015)<br>Kelly et al. (2019)<br>Petriwskyj et al. (2013)<br>Geddis-Regan et al. (2021)<br>Walsh et al. (2021)<br>Xie et al. (2018)<br>Backhaus et al. (2020) |
| ACP – Respite services and long-term place of care | “Guiding Options for Living with Dementia (GOLD)” | Helping and guide care partners make decisions to facilitate their life | ↑ understanding of respite care<br>Were able to find respite care services and to weight their preferences to help decision-making<br>Not significant: ↑ knowledge and ↓ decisional conflicts | 1 | Stirling et al. (2012) | 5 | Austin et al. (2015)<br>Davies et al. (2019)<br>Ho et al. (2021)<br>Lord et al. (2015)<br>Mattos et al. (2023) |
|  | DECIDE manual: workbook and support of a decision coach | Help care partners make decisions about long-term placement | Support for decision-making about long-term placement<br>↓ decisional conflicts | 1 | Lord et al. (2017) | 2 | Davies et al. (2019)<br>Ho et al. (2021) |
| Everyday decisions – Driving cessation | “At the Crossroads” | Education and support for care partners to address driving-related issues | ↑ confidence in managing driving cessation | 1 | Stern et al. (2008) | 2 | Davis and Ohman (2017)<br>Rapoport et al. (2012) |
| Everyday decisions – Medication management | In-person session with a nurse or social worker, and manual | Addressing seven common problems in medication management | ↑ knowledge and medication management | 1 | Lingler et al. (2016) | 1 | Gench et al. (2021) |
| eHealth –<br>Various<br>decisions | “ComputerLink” | Information, support program, and discussions with care partners and nurses | ↑ confidence<br>No effect on decision-making skills | 2 | Bass et al. (1998)<br>Brenan et al. (1995) | 1 | Hopwood et al. (2018) |
|  | “AlzMed” booklet and website | Support for medical conditions and dementia-related behaviors | ↑ confidence<br>↑ ability to manage dementia and monitor medication side effects | 1 | Zimmerman et al. (2018) | 1 | Gench et al. (2021) |
|  | “Building Better Caregiver (BBC)” internet-based workshop | Self-paced lessons, facilitator guidance, and group support | ↓ burden, depression, pain, stress | 1 | Lorig et al. (2012) | 1 | Hopwood et al. (2018) |
|  | “European eHealthMonitor project Dementia Portal (eHM-DP)” | Targeted and personalized support for care partners of persons with dementia in a home-based care setting | Informal care partners: ↑ degree of perceived support, access to support from home, confidence in health-related decision. No effect on burden and quality of life<br>Informal and professional care partners: ↑ interaction | 2 | Schaller et al. (2015, 2016) | 1 | Hopwood et al. (2018) |

**TABLE 3.**
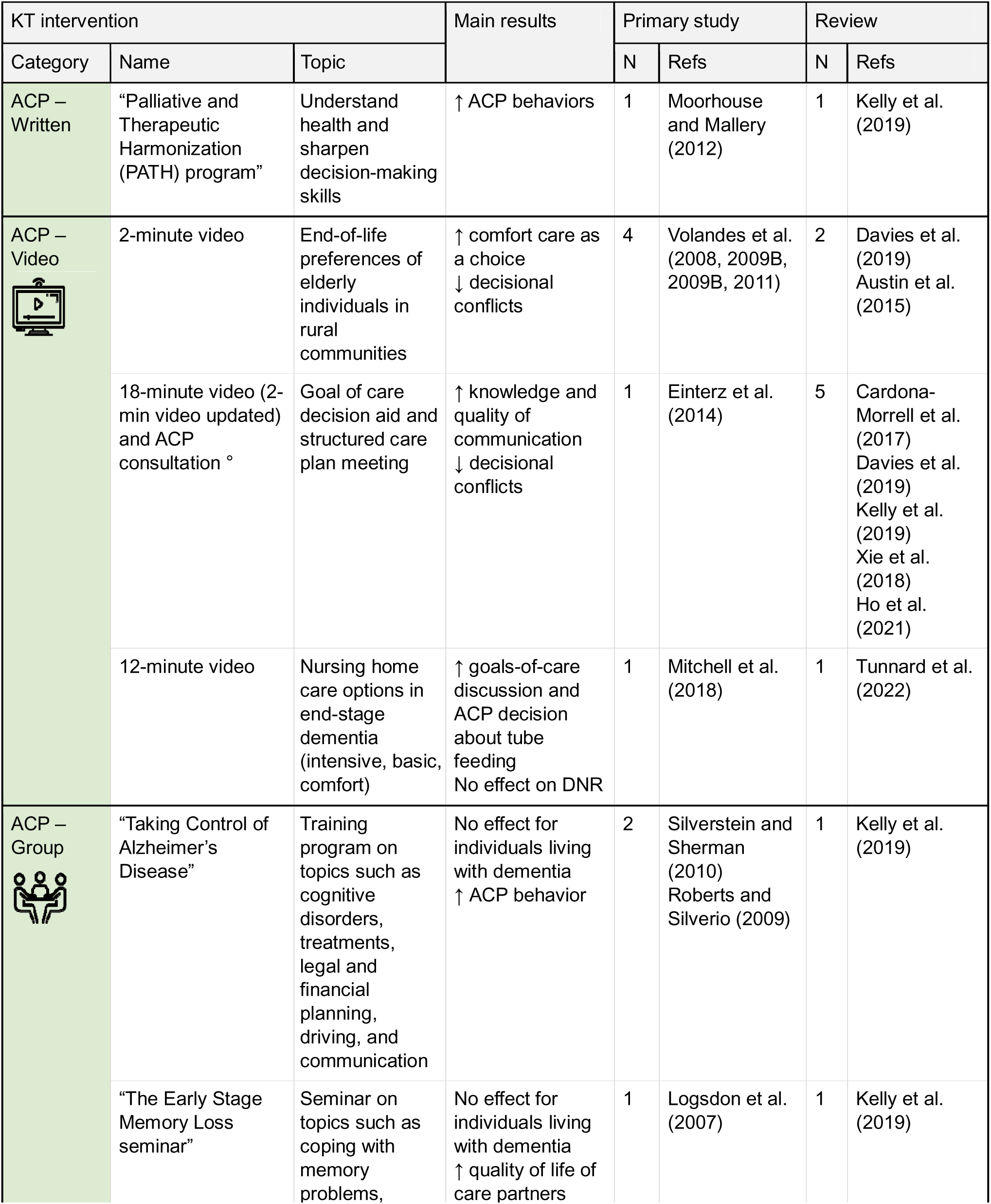
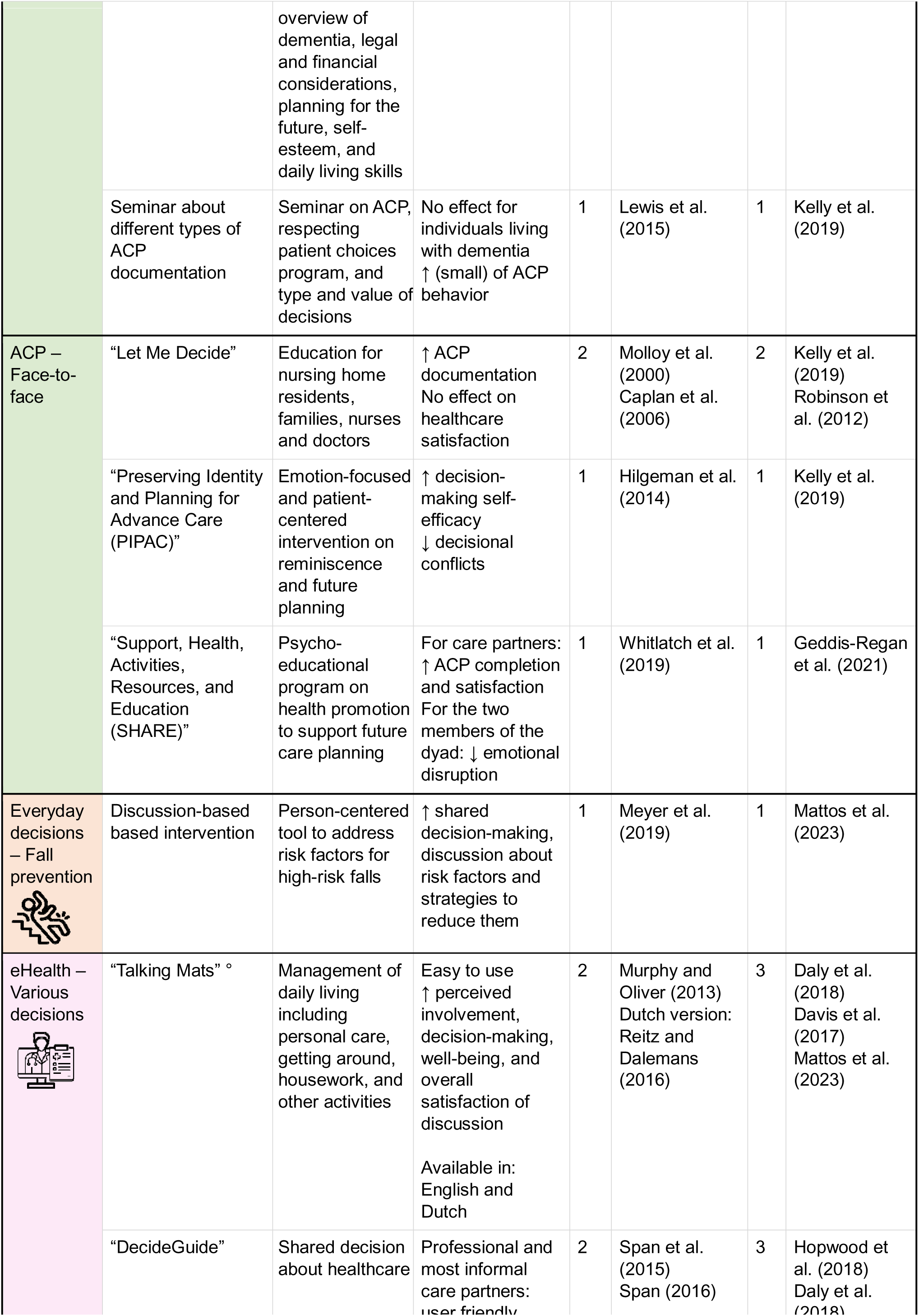
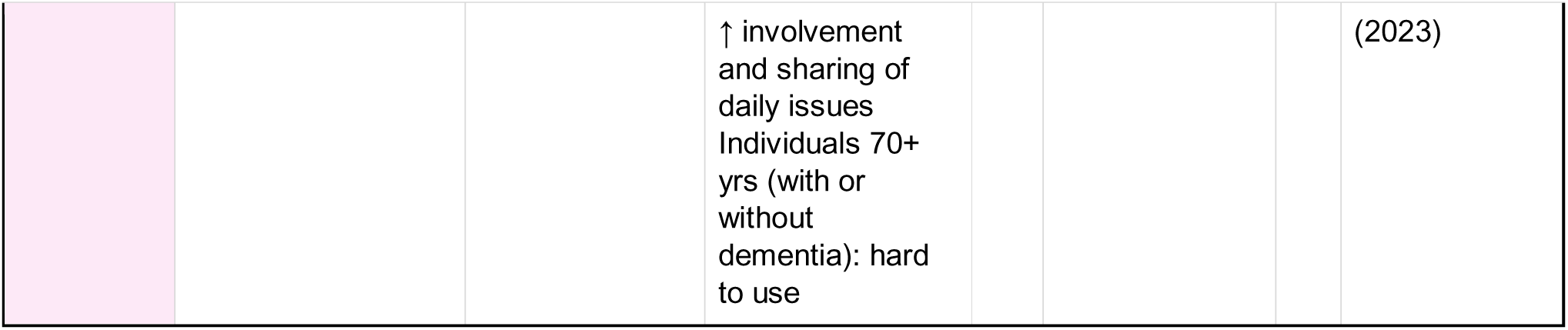
Interventions targeting dyads and triads. Note that the *“Palliative and Therapeutic Harmonization (PATH) program”* (reported by Kelly et al. (2019)) targeted older individuals without dementia. *Abbreviations*: ° multiple languages and/or cultural adaptations available; ACP, advance care planning; KT, knowledge translation.

#### 3.4.1 Advance care planning (ACP)

Of the 22 reviews included in our umbrella review [18–28,30–40], 15 reviews [18–20,23–28,31,35,36,38–40] identified 21 unique KT interventions in 40 primary articles for decisions related to end-of-life and ACP. ACP is “*the ability to enable individuals to define goals and preferences for future medical treatment and care, to discuss these goals and preferences with family and health-care providers, and to record and review these preferences if appropriate*” [42]. Examples of preferences for future medical treatment and care can include artificial feeding/hydration preference, do-not-hospitalize (DNH) orders, do-not-resuscitate (DNR) orders, or other directives such as health care power of attorney. In our umbrella review, we identified that ACP interventions (a) came in different formats, namely, written material, video decision aids, group interventions (i.e., large group sessions), and face-to-face interventions (i.e., smaller groups between individual living with dementia and/or care partner + coordinator) (sections 3.4.1.1 to 3.4.1.4), addressing general ACP topics, or (b) addressed specific topics, namely, feeding options, and respite services and long-term place of care (sections 3.4.1.5 and 3.4.1.6). ACP interventions have been summarized in the following sections (Figure 3E).

##### 3.4.1.1 ACP: Written material

Five reviews [20,26–28,39] identified two written interventions, from seven primary articles [43–49], targeting (a) care partners only, and (b) older individuals (65+ years) and their care partners. The two written interventions were found to decrease decisional conflicts and increase ACP behaviors, as further described in the following paragraphs.

Five included reviews [20,26–28,39] presented the Canadian booklet *“Comfort care at the end of life for persons with dementia – A guide for care partners”*, an informational document developed in 2005 in both French and English, targeting care partners (informal and professional) only. This booklet aimed to help care partners in their decision-making process by teaching them about dementia trajectory and palliative care approaches [20,26–28,39]. While the level of family inclusion in decision making was not evaluated [39], this written intervention (a) decreased the level of decisional conflict between care partners, and (b) did not increase the completion of the DNR order [20,28]. Upon examination of the relevant primary studies included in these reviews, we found (a) three studies featured the acceptability and usefulness of the booklet by healthcare professionals (e.g., nurse) [43–45], resulting in its adaptation in Dutch, Japanese, and Italian [46,47], and (b) one study describing an intervention combining the booklet with an ACP meeting between the informal care partner and a healthcare professional (e.g., nurse) [48].

In addition, one review [26] identified the usage of the “*Palliative and Therapeutic Harmonization (PATH) program*” targeting individuals of 65-years and older and their care partners, which combined written decision-support intervention and consultation with a healthcare professional. Specifically, this program consisted of reviewing written and online materials describing frailty and dementia using narrative stories and case vignettes, followed by three consultations with a healthcare professional (physician or nurse practitioner) [49]. This program was shown to increase ACP behavior as reported by the participants [26].

##### 3.4.1.2 ACP: Video decision aids

Nine reviews [20,23–27,31,35,38] identified four video decision aid interventions, from ten primary articles [50–59], targeting both individuals with dementia and their care partners, and individuals with advanced dementia only. Overall, video decision aid interventions were found to positively impact individuals with dementia and their care partners, by increasing knowledge and goals-of-care decision making, and by decreasing decisional conflicts. More details about the video interventions are provided below.

Specifically, two reviews [24,31] identified a two-minute video targeting individuals with dementia and their care partners, which contained an audio description of advanced dementia [50–52,57]. Compared to a narrative/descriptive format (i.e., without visual format), this video significantly increased comfort care as a choice, and decreased the level of decisional conflict between individuals over 65-years-old and their care partners [24,31]. Later, Einterz et al. [58] created a longer version (18-minute video) of the above-mentioned two-minute video and combined it with an ACP consultation [24–27,38,58], which increased knowledge and decreased decisional conflict between individuals with moderate-to-severe dementia and their care partners (both family members and healthcare professionals) [24,25,38]. This intervention (i.e., the 18-minute video combined with an ACP consultation) was more recently evaluated in a randomized clinical trial [54] and similar beneficial effects were reported [20,24,27,35]. Specifically, it was shown to be acceptable for care partners by increasing (a) knowledge, (b) quality of communication [24,27], and (c) decreasing decisional conflicts both with the individual living with dementia [24,35], and with health professionals (e.g., clinicians) [20,24]. However, the lower level of conflict between care partners and health professionals was considered to be of low certainty in a subset analysis by Walsh et al. [35] for dyads in a late-dementia stage.

Finally, one review [23] identified two additional video decision-support interventions: (a) one 12-minute video targeting both individuals with advanced dementia and their care partners [55], and (b) five 6-to-10-minute videos targeting only individuals with advanced dementia only [56]. While these two video interventions were found to increase goals-of-care discussions and ACP decision about tube feeding, no impact was shown on DNH decision making [23].

##### 3.4.1.3 ACP: Group interventions

Two reviews [26,28] identified five group interventions, from six primary studies [60–65], targeting (a) family care partners, and (b) both individuals living with dementia and their care partners. The two reviews concluded that while the interventions did not impact outcomes for individuals living with dementia, they were beneficial for care partners and seemed to increase ACP interest as inferred from self-reported measures [26,28]. Details about the five interventions are provided below.

Group interventions targeting family care partners were: (a) the *“Legal-Financial Planning Workshop”* [64], and (b) an educational group intervention (including presentations and printed materials) [60] about dementia trajectory, life support, tube-feeding, and ACP for African American care partners, which increased their knowledge and decision-making self-efficacy regarding end-of-life care [26,28].

Of the three group interventions targeting individuals living with dementia and their care partners identified by one review [26], two were sponsored by the Alzheimer’s Association: (a) the “*Taking Control of Alzheimer’s Disease*”, a 4-session intervention including ACP discussions at early-stage dementia related to finances, driving, and legal issues [61,62]; and (b) “*The Early Stage Memory Loss seminar”*, a similar intervention including ACP and financial planning [63]. The third intervention consisted of a seminar about different types of ACP documentation [65].

##### 3.4.1.4 ACP: Face-to-face interventions

Seven included reviews [18–20,26,28,29,35] identified seven unique face-to-face interventions/consultations, from eight primary studies [66–73], regarding end-of-life decisions. These interventions were targeted at (a) care partners of individuals living with dementia, or (b) both individuals living with dementia and their care partners. Overall, the reviews agreed that face-to-face interventions increased the level of comfort for decision making and ACP completion. Findings relative to these face-to-face interventions are described in more details below.

Regarding interventions targeted at care partners of individuals living with dementia, one review [40] identified one face-to-face intervention targeting Chinese family care partners of older people living with dementia. This intervention involved presenting hypothetical scenarios featuring critical illness or irreversible coma to care partners, followed by information on possible life sustaining treatment outcomes [40,73]. The intervention was found to slightly improve the level of comfort for decision making in the case of irreversible coma compared to critical illness, suggesting that the context (e.g., coma or critical illness) affected the decisions [40,73]. Five additional reviews [18,20,26,28,35] identified three interventions involving a specialized palliative care team targeting care partners of individuals living with advanced dementia [68–70]: (a) a structured palliative care consultation that included the opportunity to complete ACP followed by phone support by a palliative care nurse practitioner, with care partners receiving the booklet “*Advanced Dementia: A Guide for Families”* [20,35,68], (b) a palliative consult intervention delivered by a physician and a palliative care social worker [26,28,69], and (c) a nurse-delivered adapted version of the “*UK National Health Service Preferred Priorities of Care*” [70]. While the first two of the above-mentioned interventions reported positive effects, such as increased completion of ACP documentation and improved care partners decision making [20,26,28,35], the third intervention, in contrast, was shown to increase decisional conflict of care partners, with few of them completing ACP documentation [18,70].

Three reviews identified interventions targeting both individuals living with dementia and their care partners. Of these, two reviews [19,26] mentioned the “*Let Me Decide”* advanced directive program/intervention, which trained health professionals to counsel individuals living with dementia and their care partners about ACP [66,67]. The intervention was found to increase ACP documentation but did not change healthcare satisfaction [19,26]. Targeting individuals living with early-stage dementia and their care partners, two reviews [20,26] identified two face-to-face interventions [71,72]: (a) the “*Preserving Identity and Planning for Advance Care (PIPAC)”* intervention [26,71] consisting of reminiscence and ACP sessions, and (b) the “*Support, Health, Activities, Resources, and Education (SHARE)*” psychoeducational program [20,72], an intervention including counseling on ACP. Both interventions (i.e., PIPAC and SHARE) were found to have positive effects, with PIPAC reporting decreased decisional conflict and improved decision-making self-efficacy [26], and with SHARE demonstrating increased ACP completion and satisfaction for care partners only, and decreased emotional disruption for both members of the dyad (i.e., individuals living with early-stage dementia and their care partners) [20].

##### 3.4.1.5 ACP: Feeding options

Twelve reviews [18,20,24–27,31,34,35,38–40] identified one ACP intervention for decision making about feeding options, from eight primary studies [74–81]. Five reviews [18,24,25,27,34] identified the original intervention by Mitchell et al. [79], specifically, the audiovisual intervention “*Making Choices: long-term tube feeding placement in elderly patients”* and personal worksheet. While this decision aid was initially targeted at care partners of older individuals (65+ years of age) [79], three more recent updates [20,24–27,31,34,35,38–40] of this intervention targeted: (a) dyads of individuals living with advanced dementia and their care partners [76,77], and (b) only care partners of individuals living with advanced dementia [75,80]. All twelve reviews noted that both the primary intervention study and its updates reported decreased decisional conflicts and expectations about tube feeding benefits, as well as increased goals-of-care, and knowledge and frequency/quality of communication [18,20,24–27,31,34,35,38–40]. Despite this, two reviews [20,35] found limitations in the updated versions, with limited degree of change in knowledge and decisional conflict, especially for care partners of individuals living with advanced dementia. However, due to the overall positive impact of this intervention, two reviews [24,34] noted that it was adapted into the “*International Patient Decision Aid Standards (IPDAS)* statement” by Elwyn et al. [81] (with up to six versions in different languages, such as Spanish and Chinese, translated from the original English version), as well as translated to other languages [34] including Portuguese [74] and Japanese [78].

##### 3.4.1.6 ACP: Respite services and long-term place of care

Five reviews [18,24,27,31,36] identified two interventions, from two primary studies [82,83], targeting only care partners for decision making about respite services and long-term place of care. Specifically, all five reviews [18,24,27,31,36] identified the “*Guiding Options for Living with Dementia (GOLD)*”, a decision aid book from Australia targeting dementia care partners [83]. This intervention allowed care partners to (a) understand respite care through synthesized information and vignettes about care partners experiences, (b) find community respite care services, and (c) weigh their preferences to help make decisions [83]. While the GOLD intervention was found to be useful and relevant by care partners [24], only a non-significant trend towards lower decisional conflicts and higher knowledge was observed [18,24]. In addition, two reviews [24,27] identified the DECIDE manual [82], that supported care partners in their choice of long-term care placement through the completion of a workbook with the support of a decision coach. The DECIDE manual was found to reduce decisional conflicts of care partners [24,27].

#### 3.4.2 Everyday decision making

Of the 22 reviews included in our umbrella review [18–28,30–40], four reviews [27,30,32,36,37] identified four unique KT interventions in four primary articles for decisions related to everyday decision making. Everyday decision making is the ability to solve problems, and make decisions about everyday situations such as choosing what to wear or what to do [33]. Within these included articles, we identified three everyday decision-making topics, specifically driving cessation, fall prevention, and medication management. KT interventions about everyday decision making have been summarized in the following sections (Figure 3F).

##### 3.4.2.1 Driving cessation

Three reviews [27,32,37] identified two interventions, from two primary studies [84,85], for decision making about driving cessation targeting either care partners or individuals living with dementia, and noted beneficial impact. Specifically, two reviews [32,37] identified “*At the Crossroads”* intervention [84], a psycho-educational group intervention for care partners of individuals living with MCI, AD, or related dementia. The intervention increased care partners’ confidence in managing driving cessation of their relatives living with dementia [32,37]. One review [27] identified the “*Driving with Dementia Decision Aid (DDDA)”* booklet [85], which targeted drivers living with dementia. The DDDA reduced decisional conflict and improved knowledge of individuals with dementia regarding driving cessation [27].

##### 3.4.2.2 Fall prevention

One review [36] identified one intervention for decision making about fall prevention from one primary study [86]. The discussion-based intervention was useful for supporting shared decision making by individuals living with dementia and their care partners, by discussing fall risk factors and strategies to reduce them, as well as their advantages and disadvantages [36].

##### 3.4.2.3 Medication management

One review [30] identified an educational intervention for care partners, from one primary study [87], for decision making about medication management. In-person sessions with a nurse or social worker were guided by a manual addressing seven areas of medication management: care partners responsibilities, common problems in medication administration/taking, preventing medication errors, talking with health care providers about medications, community resources, contingency planning, and changes in medication taking. The intervention improved care partners knowledge and medication management [30].

#### 3.4.3 eHealth interventions

Finally, six reviews [21,22,27,30,33,36] identified seven interventions, from eleven primary studies [88–98], addressing multiple decision topics related to living with dementia. These interventions were eHealth interventions, defined as “*the use of information and communication technologies for health*” by the World Health Organization [99]. eHealth interventions offer a distinct delivery format that is flexible, scalable, and easily personalized, while also allowing individuals to engage at their own pace [100]. These seven eHealth interventions targeted (a) care partners only (N=4 interventions; informal only, or informal and professional), (b) individuals living with MCI only (N=1), (c) both individuals living with dementia and their care partners (N=1), and (d) triads of individuals living with dementia, family care partners, and professionals (N=1). It was found that eHealth interventions for decision making about current or future matters had beneficial effects, especially for care partners (both informal and professional). Findings relative to these eHealth interventions are described in more details below.

Four reviews targeted (a) family care partners of AD individuals, and (b) both family and healthcare professionals. Specifically, one review [21] identified “*ComputerLink*” [88,89], a digital-based tool with three main functions: (a) information delivery through an electronic encyclopedia, (b) decision support program that used prioritization questions, and (c) communication with other care partners and nurses using a forum, private messages, and a Q&A module. This intervention had a positive impact on decisional family care partners’ confidence, although it did not change decision-making skill and the decision support module may have been used sub-optimally [21]. In addition, two reviews [21,30] identified two other interventions for care partners: (a) Gench et al. [30] identified “*AlzMed*”, a booklet and website [98] that significantly improved care partners’ confidence and ability to manage dementia and monitor medication side effects, and (b) Hopwood et al. [21] identified the “*Building Better Caregiver (BBC)*”, an internet-based, skills-enhancement workshop [90] that significantly reduced care partners’ burden, depression, pain, and stress. Finally, one review [21] described an intervention for informal care partners of individuals living with dementia and healthcare professionals, the decision aid “*European eHealthMonitor project Dementia Portal (eHM-DP)*” [91,92]. Family care partners reported (a) high degree of perceived support by the eHM-DP from individualized information acquisition, access to support from home, and empowerment in health-related decisions, and (b) improved interactions between informal care partners and healthcare professionals [21]. However, it did not improve care partners’ burden and quality of life over the study period [92].

One review [27] identified a promising web-based intervention for decision making for older individuals living with MCI (60+ years) based on the personal values and preferences of the users [97]. In addition, three reviews [22,33,36] identified the “*Talking Mats*” [95], a picture-based communication framework for individuals with dementia and their family care partners. The “*Talking Mats*” (both the English [95] and Dutch version [101]) was reported to be easy to use and to increase (a) the perceived involvement of both members of the dyads, (b) decision making, (c) well-being, and (d) overall satisfaction of the discussion [22,33,36]. However Mattos et al. [36] reported that despite a request for adding pictures and decreasing complexity of interventions using decision boxes, the overall feeling was positive regarding the decision aid for assistance in decision making [96].

Finally, three reviews [21,22,36] identified an intervention for triads (individuals living with dementia, informal care partners, case managers – i.e., relatively new term for individuals helping dyads of individuals with dementia and their care partners). Specifically, they described “*DecideGuide*”, an interactive web-based intervention for eight dementia-related life domains [93,94]. Case managers and most family care partners found this intervention to be user friendly, and (a) appreciated this tool (especially the chat function), (b) felt more involved, and (c) were able to share more information about daily issues [21,22,36]. However, it was noted that older adults (70+ years), with or without dementia, found it harder to use [21,22].

## 4. DISCUSSION

In our umbrella review, we assessed existing KT interventions aimed at helping individuals living with dementia and their care partners make decisions about current or future matters. Since the involvement of individuals living with dementia was found to be limited and difficult, due to their lack of understanding of topics that require decision making, it is crucial to develop and offer interventions to increase their knowledge and help them being involved [11,26]. Specifically, we presented a total of 32 KT interventions for decision making from 22 reviews. Of these 32 interventions, 21 (66%) were ACP interventions, highlighting the importance of ACP as a critical component for decision making about current or future medical treatment or care. Based on the findings from prior systematic reviews, which found that written, video, and face-to-face KT interventions for ACP generally increased knowledge and reduced decisional conflict, we recommend that ACP interventions be made more available, both in terms of marketing and knowledge, to help older individuals (with or without dementia) and their care partners (informal or professional).

### 4.1 Targeted population

#### Care partners

Out of the 32 KT interventions, 16 (50%) targeted care partners only (Table 2), with the three most cited being ACP interventions, namely: (a) the Canadian booklet *“Comfort care at the end of life for persons with dementia – A guide for caregivers”* [20,26–28,39,43–48], (b) the audiovisual intervention *“Making Choices: long-term tube feeding placement in elderly patients”* and worksheet (including the three updates an three cultural adaptations) [18,20,24–27,31,34,35,38–40,74–81], and (c) the Australian book *“Guiding Options for Living with Dementia (GOLD)”* [18,24,27,31,36,83]. Overall, all KT interventions targeted at care partners were found to have beneficial effects such as (a) increased knowledge (N=5), (b) increased confidence (N=4), and (c) decreased decisional conflicts (N=4). Only one KT intervention targeted at care partners, namely the “*UK National Health Service Preferred Priorities of Care*”, had a negative effect with increased decisional conflicts reported [18,70]. This effect was negative due to the fact that many care partners were resistant to make decisions about hypothetical future scenarios, highlighting the importance of being prepared and enthusiastic about such interventions.

#### Individuals living with dementia and care partners

A total of 13 (41%) KT interventions were targeted at both individuals living with dementia and their care partners (N=12 dyads; N=1 triad; Table 3). The most cited intervention was an 18-minute video combined with an ACP consultation [24–27,38,58]. Targeting individuals living with dementia only (Table 4), the three (9%) KT interventions reported in our review were all found to have beneficial impacts on individuals living with MCI to advanced dementia [23,27,56,85,97]. Overall, across all 32 KT interventions, while the interventions were shown to have positive effects, the involvement of individuals living with dementia was low, and mostly at early stages of dementia. We therefore recommend more KT interventions targeted at individuals living with dementia.

**TABLE 4.**
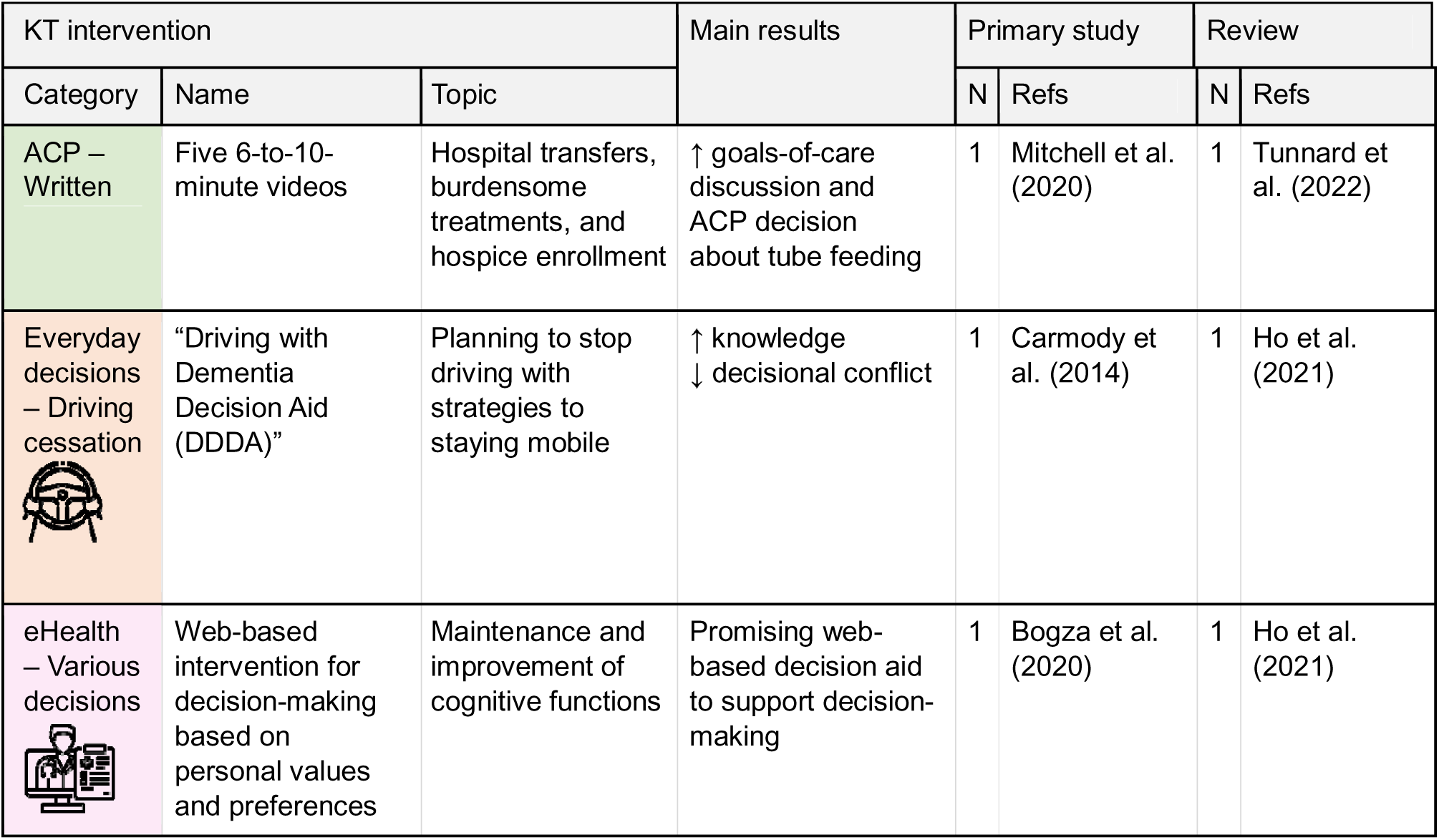
Interventions targeting individuals living with dementia. *Abbreviations*: ACP, advance care planning; KT, knowledge translation.

| KT intervention |  |  | Main results | Primary study |  | Review |  |
| --- | --- | --- | --- | --- | --- | --- | --- |
| Category | Name | Topic |  | N | Refs | N | Refs |
| ACP – Written | Five 6-to-10-minute videos | Hospital transfers, burdensome treatments, and hospice enrollment | ↑ goals-of-care discussion and ACP decision about tube feeding | 1 | Mitchell et al. (2020) | 1 | Tunnard et al. (2022) |
| Everyday decisions – Driving cessation | “Driving with Dementia Decision Aid (DDDA)” | Planning to stop driving with strategies to staying mobile | ↑ knowledge<br>↓ decisional conflict | 1 | Carmody et al. (2014) | 1 | Ho et al. (2021) |
| eHealth – Various decisions | Web-based intervention for decision-making based on personal values and preferences | Maintenance and improvement of cognitive functions | Promising web-based decision aid to support decision-making | 1 | Bogza et al. (2020) | 1 | Ho et al. (2021) |

### 4.2 Diversity of targeted population

Overall, six KT interventions were culturally and/or linguistically adapted (N=5 ACP, N=1 eHealth; Tables 2, 3). Specifically, we found that three ACP interventions all originally published in English, were later translated in several languages such as Dutch, Japanese, Portuguese, French, Italian, Spanish, and Chinese, namely: (a) *“Comfort care at the end of life for persons with dementia – A guide for caregivers”* [20,26–28,39,43–48], (b) *“Making Choices: long-term tube feeding placement in elderly patients”* and worksheet [18,24,25,27,34,74–81], and (c) the 18-minute video and consultation by Einterz et al. [24–27,38,58]. Furthermore, while ethnicity was not reported by the majority of studies (91%), two ACP interventions targeted African Americans (both individuals living with dementia and care partners) [24–28,38,58,60], and one ACP intervention targeted Chinese care partners specifically [40,73]. In addition, one eHealth intervention, specifically, the “*Talking Mats*”, was also translated from English to Dutch [22,33,36,95,101].

We recommend that more English KT interventions be adapted to different languages and cultures, to reach a larger part of the population and address a broader range of issues individuals living with dementia and/or their care partners may face. Since our search was performed only in English, it is possible that KT intervention reviews in other languages may have been missed. However, this scenario is unlikely given that findings from our pilot search demonstrated that expanding to additional languages did not yield substantially more review articles.

### 4.3 Limitations

Three main limitations were identified. First, the heterogeneity of format delivery (e.g., face-to-face interventions, videos) and of decision-making outcome measures (e.g., level of knowledge, decisional conflicts), complicated the comparison of different interventions. We therefore recommend standardization of outcome measures to enable more reliable comparison of KT intervention effectiveness, as this would make it easier to compare outcomes across studies, including pooling data for meta-analysis. Scales have been developed to assess decision making, including the “*Decisional Conflict Scale*” (https://decisionaid.ohri.ca/eval_dcs.html; [102]) and the “*Satisfaction with Decision scale*” [103]. Achieving greater consensus on decision-making measurement has been the focus of organizations such as the “*International Patient Decision Aids Standards*” (IPDAS; https://decisionaid.ohri.ca/IPDAS/; [81]). However, while the diversity of format delivery limits generalization and comparison of different interventions, it could facilitate implementation of decision-making interventions and allow the delivery to be better tailored to specific settings. Secondly, many interventions identified were tested in small pilot studies, which limited the quality of evidence presented, thus, results should be interpreted with caution. In addition, since our review is an umbrella review, it should be noted that all developed interventions may not have been retrieved since we depended on published reviews, which themselves were dependent on the quality of primary articles. Quality assessment of primary studies was not always carried out adequately by the retrieved reviews. Specifically, five reviews did not conduct quality assessment, and of the 17 that did, seven were not done independently by more than one reviewer. Another limitation of the included reviews was that only two studies addressed the likelihood of publication bias. Despite these limitations, our umbrella review provides a good portrait of topics addressed by KT interventions, available in the literature thus far.

### 4.4 Clinical implications

The findings from this umbrella review address an important gap in the care of individuals living with dementia, who face difficult decisions as a result of their illness, including ones about safety, medical care, and, eventually, end-of-life care. Given the effects of dementia on reasoning and executive function, care partners must necessarily share some of, or even all of, the burden of decision making. This can cause stress and decrease quality of life for both care partners and individuals living with dementia, particularly when their values are incongruent [104]. As this review shows, there are KT interventions that can decrease stress and decisional conflict, potentially improving quality of life. Although more research is needed, we suspect that standardized, validated decision-making interventions are probably under-utilized and/or less known in routine clinical care. Barriers to adoption include lack of awareness of these interventions and their impact, lack of access to the tools, and lack of resources, including staff time, to provide interventions for decision support, particularly for face-to-face interventions. However, we found evidence that thirteen (41%) of the 32 KT interventions are currently available on the internet (Supplementary Material S4), and could be rapidly adopted into practice by interested clinicians. Examples of KT intervention freely accessible online include the *“Comfort care at the end of life for persons with dementia – A guide for care partners”*, the *“Palliative and Therapeutic Harmonization (PATH) program”*, and the “*Talking Mats”*. Further development of eHealth interventions might allow greater access to decision-making support at a reasonable cost and improve access to care in communities without dementia specialists, although some have been found to be harder to use for older individuals. Finally, we recommend that these KT interventions be brought together as a toolkit for healthcare system navigators to introduce to both individuals living with dementia and their care partners. Health systems should therefore consider investing more resources into decision support.

### 4.5 Future directions

Future research should focus on (a) the validation of developed KT interventions in diverse cohorts, ideally using a multicenter clinical trial design-based approach, (b) the development and/or adaptation of KT interventions that specifically target individuals living with MCI or dementia, ideally at earlier stages of the disease to enable their involvement in the decision-making process, and (c) the dissemination of KT interventions to enable more widespread adoption of the tools. In addition, with the advent of new eHealth and artificial intelligence tools such as large language models, it will be increasingly feasible to incorporate and use these models to streamline healthcare communication and education [105]. It is important that the interventions are tested robustly in different populations and with different practitioners (if applicable) to ensure the feasibility of the interventions. Such studies could also enable higher levels of dissemination and increase their use in the clinic. It is possible that cultural adaptations may be needed for specific groups and thus incorporating feedback from a diverse study population could increase efficacy and update.

## 5. CONCLUSION

In conclusion, this umbrella review addresses a critical gap by uniquely synthesizing existing KT interventions that support decision making in dementia care. Our umbrella review provides: (a) a structured stratification of interventions by targeted knowledge users, including individuals living with dementia and informal care partners; (b) a dedicated section on clinical and implementation implications, specifically addressing the needs of healthcare professionals and researchers; and (c) the identification of critical evidence gaps, such as the lack of comparative studies and standardized outcome measures, to guide future research. We recommend continued implementation and dissemination of proven tools, along with efforts to raise awareness. Further studies are also needed to address different stages of dementia and to include more culturally diverse populations.

## Data Availability

All data produced in the present work are contained in the manuscript

## AUTHOR CONTRIBUTIONS

A.B., M.B., and E.E.S. designed the study. The search strategy was developed by M.B., E.L.M., E.E.S., and A.B. The screening process was performed independently by M.B., F.E.D., S.A., S.B., W.B., P.F.B., K.S.G., J.P., and E.E.S. Full-text review of included publications was done by M.B., F.E.D., and E.E.S. Clinical appraisal and data extraction were done by M.B. and F.E.D. First draft was written by M.B., F.E.D., J.P., E.E.S., and A.B. All authors edited and approved the final manuscript. All authors meet the ICMJE criteria for authorship.

## ACKNOWLEDGEMENTS

Not applicable.

## CONSENT STATEMENT

We confirm that consent was not necessary for this work.

