## Supplementary Material S1 for "Supporting decision making for individuals living with dementia and their care partners with knowledge translation: an umbrella review"

To enable PROSPERO to focus on COVID-19 submissions, this registration record has undergone basic automated checks for eligibility and is published exactly as submitted. PROSPERO has never provided peer review, and usual checking by the PROSPERO team does not endorse content. Therefore, automatically published records should be treated as any other PROSPERO registration. Further detail is provided here.

### Citation

Marie Biard, Flavie Detcheverry, Patrick Bloniasz, Sandi Azab, Sara Becker, William Betzner, Karl S. Grewal, Jolene Phelps, Eric E. Smith, AmanPreet Badhwar. Supporting decision-making for people living with dementia and their caregivers with knowledge translation : an umbrella review. PROSPERO 2023 CRD42023414419 Available from: https://www.crd.york.ac.uk/prospero/display_record.php?ID=CRD42023414419

### Review question

1. What types of knowledge translation strategies exist for people living with dementia and caregivers to improve informed decision-making ?

2. What are the outcomes of knowledge translation strategies for people living with dementia and caregivers ?

3. What gaps exist in knowledge translation about decision-making for persons living with dementia and caregivers ?

### Searches

We conducted a search on January 7, 2023 using 4 electronic databases on Ovid: MEDLINE, APA PsycINFO, EMBASE and Cochrane database of systematic reviews. The search wasn't limited to a publication period. Language was restricted to English. No unpublished systematic reviews will be included.

### Types of study to be included

Review article that used a defined, reproducible search strategy; systematically applied inclusion and exclusion criteria; and a synthesis of the returned evidence, which may be qualitative. These reviews are usually self identified as: "Systematic review", "Meta-analysis", "Rapid review", "Umbrella review" and "Scoping review". Pre-print reviews or review protocols will be excluded.

### Condition or domain being studied

Dementia

### Participants/population

Inclusion:

1. Persons with dementia, or with these dementia subtypes (i.e., dementia-related knowledge translation):

- Dementia or cognitive impairment in general

- VCID/vascular dementia

- Alzheimer's disease

- Mixed dementia

2. Informal caregivers of the above population.

Exclusion:

Knowledge translation intervention directed at:

1. Persons with other dementia subtypes (e.g., Lewy Body dementia, Parkinson related dementia)

2. Persons with other illnesses (e.g., stroke) or to older persons in general

3. Health professionals only

4. Professional caregivers only

### Intervention(s), exposure(s)

Knowledge translation studies about strategies/interventions that support persons living with dementia and caregivers with decision making.

### Comparator(s)/control

Any types of comparators used in original systematic reviews.

### Main outcome(s)

Measures used in original systematic reviews e.g., increase in knowledge about dementia/dementia care and decision-making skills, satisfaction of quality of care, confidence.

### Additional outcome(s)

None.

### Data extraction (selection and coding)

All retrieved records will be imported in the Covidence platform. Nine independent reviewers participating in the Vascular Training (VAST) Platform collaboration initiative will screen titles and abstracts against inclusion/exclusion criteria. Every record will be screened by the principal author. Conflicts will be resolved by two reviews by discussion, with the help of a third reviewer if needed. Full-text of included records will be reviewed by four reviewers, and conflicts will be resolved in a similar manner.

For data extraction, two independent reviewers per article will use a modified version of the JBI Data Extraction Tool (Aromataris et al., 2015). Specifically, the following elements will be extracted in a standardized manner: author, year, country, objective(s), participants characteristics, setting, interventions, databases searched, date range of included studies, basic information about primary studies, appraisal instrument and rating, type of review, outcome(s), results and other comments. Reviewers will have done a pilot data extraction practice beforehand.

### Risk of bias (quality) assessment

Two reviewers will independently conduct the quality assessment of selected systematic reviews using the JBI Critical

Appraisal Checklist for Systematic Reviews and Research Syntheses (Aromataris et al., 2015). Briefly, the checklist includes eleven questions, nine of which assess the validity of the study. The last two questions assess the relevance of the recommendations and gaps identified. Reviewers will have done a pilot quality assessment practice beforehand. Conflicts in quality assessment will be resolved by discussion, with the help of a third reviewer if needed.

### Strategy for data synthesis

The extracted data will be presented in various summary/findings tables as well as reported in a narrative format. A thematic analysis will be performed to categorize knowledge translation interventions (e.g. type of use).

### Analysis of subgroups or subsets

If sufficient data are available, knowledge translation interventions specifically directed at persons living with Azlheimer's disease or Vascular Cognitive Impairment and Dementia (VCID) may be addressed.

### Contact details for further information

AmanPreet Badhwar

### Organisational affiliation of the review

Centre de Recherche Institut Universitaire de Gériatrie de Montreal (CRIUGM), Université de Montréal

http://www.criugm.qc.ca/; https://pharmacologie-physiologie.umontreal.ca/

### Review team members and their organisational affiliations

Ms Marie Biard. CRIUGM & Université de Montréal

Ms Flavie Detcheverry. CRIUGM & Université de Montréal

Mr Patrick Bloniasz. Boston University

Dr Sandi Azab. McMaster University

Dr Sara Becker. University of Calgary

Mr William Betzner. University of Calgary

Mr Karl S. Grewal. University of Saskatchewan

Dr Jolene Phelps. University of Calgary

Dr Eric E. Smith. University of Calgary

Dr AmanPreet Badhwar. CRIUGM & Université de Montréal

### Type and method of review

Review of reviews, Systematic review

### Anticipated or actual start date

01 February 2023

### Anticipated completion date

31 May 2023

### Funding sources/sponsors

Canadian Institutes of Health Research (RT0 179993)

### Conflicts of interest

### Language

English

### Country

Canada

### Stage of review

Review Ongoing

### Subject index terms status

Subject indexing assigned by CRD

### Subject index terms

Caregivers; Decision Making; Dementia; Humans; Translational Science, Biomedical

### Date of registration in PROSPERO

16 April 2023

### Date of first submission

05 April 2023

### Stage of review at time of this submission

| **Stage** | **Started** | **Completed** |
|---|---|---|
| Preliminary searches | Yes | Yes |
| Piloting of the study selection process | Yes | Yes |
| Formal screening of search results against eligibility criteria | Yes | No |
| Data extraction | No | No |
| Risk of bias (quality) assessment | No | No |
| Data analysis | No | No |

*The record owner confirms that the information they have supplied for this submission is accurate and complete and they understand that deliberate provision of inaccurate information or omission of data may be construed as scientific misconduct.*

*The record owner confirms that they will update the status of the review when it is completed and will add publication details in due course.*

### Versions

16 April 2023

16 April 2023
