## Supplementary Material S2 for "Supporting decision making for individuals living with dementia and their care partners with knowledge translation: an umbrella review"

| | | |
|---|---|---|
| KT | 1 | exp Translational Science, Biomedical/ |
| | 2 | (("knowledge" or "research" or "evidence" or "guideline*" or "information") adj2 ("translation" or "implementation" or "utili*" or "diffusion" or "management" or "use" or "exchange" or "mobili*" or "dissemination" or "transfer" or "transmission" or "uptake")).tw. |
| | 3 | KT.tw. |
| | 4 | ("integrated knowledge translation" or "integrated KT").tw. |
| | 5 | knowledge to action.tw. |
| | 6 | bench to bedside.tw. |
| | 7 | implementation science.tw. |
| | 8 | (intervention* or strateg* or tool* or aid*).tw. |
| | **9** | **or/1-8** |
| Decision making | 10 | exp Decision Making/ |
| | 11 | exp Decision Support Techniques/ |
| | 12 | exp Decision Support Systems, Clinical/ |
| | 13 | (decision* or deciding or choice*).tw. |
| | **14** | **or/10-13** |
| Knowledge users | 15 | exp Patients/ |
| | 16 | caregivers/ |
| | 17 | exp Family/ |
| | 18 | caregiving dyad.tw. |
| | 19 | ("patient*" or "consumer*" or "user*" or "carer*" or "caregiver*" or "client*" or "famil*").tw. |
| | **20** | **or/15-19** |
| Dementia | 21 | exp Dementia/ |
| | 22 | dementia.tw. |
| | 23 | exp Dementia, Vascular/ |
| | 24 | vascular cognitive impairment.tw. |
| | 25 | vascular cognitive impairment and dementia.tw. |
| | 26 | VCID.tw. |
| | 27 | cerebral amyloid angiopathy.tw. |
| | 28 | post-stroke cognitive impairment.tw. |
| | 29 | post-stroke dementia.tw. |
| | 30 | major vascular neurocognitive disorder.tw. |
| | 31 | minor vascular neurocognitive disorder.tw. |
| | 32 | acute onset vascular dementia.tw. |
| | 33 | arteriosclerotic dementia.tw. |
| | 34 | binswanger's disease.tw. |
| | 35 | subcortical leukoencephalopathy.tw. |
| | 36 | subcortical arteriosclerotic encephalopathy.tw. |
| | 37 | subcortical vascular dementia.tw. |
| | 38 | subcortical ischemic vascular dementia.tw. |
| | 39 | multi-infarct dementia.tw. |
| | 40 | cerebral autosomal dominant arteriopathy with subcortical infarcts and leukoencephalopathy.tw. |
| | 41 | CADASIL.tw. |
| | 42 | cerebral autosomal recessive arteriopathy with subcortical infarcts and leukoencephalopathy.tw. |
| | 43 | CARASIL.tw. |
| | **44** | **or/21-43** |
| Reviews | 45 | Review/ or "Systematic Review"/ or "Review Literature as Topic"/ |
| | 46 | ("Review" or "Systematic Review" or "Review Literature as Topic").af. |
| | 47 | Meta-Analysis/ |
| | 48 | Meta-Analysis.af. |
| | **49** | **or/45-48** |
| | **50** | **and/9,14,20,44,49** |
| | **51** | **limit 50 to (english language and humans)** |
