## Supplementary Material S4 for "Supporting decision making for individuals living with dementia and their care partners with knowledge translation: an umbrella review"

**SUPPLEMENTARY TABLE S4. Availability of intervention online**

| KT intervention name | Targeted population | Primary refs | Review refs | Available online | Link to intervention |
| --- | --- | --- | --- | --- | --- |
| "Comfort care at the end of life for persons with dementia – A guide for caregivers” | Care partners | Arcand et al. (2013)  van der Steen et al. (2011A, 2011B, 2012 2013)  Brazil et al. (2018) | Kelly et al. (2019)  Ho et al. (2021)  Bryant et al. (2019)  Backhaus et al. (2020)  Geddis-Regan et al. (2021) | Yes | <https://caregiversns.org/images/uploads/all/ComfortCareBooklet_EN.pdf> |
| “Legal-Financial Planning Workshop” | Family care partners | Pratt et al. (1989) | Bryant et al. (2019)  Kelly et al. (2019) | Yes | <https://www.caregiveroc.org/event-details/virtual-legal-financial-planning-workshop-3> |
| Educational group intervention | African American family care partners | Bonner et al. (2014) | Bryant et al. (2019)  Kelly et al. (2019) | No |  |
| Hypothetical scenarios (critical illness or irreversible coma) | Chinese family care partners of older individuals living with dementia | Kwok et al. (2007) | Petriwskyj et al. (2013) | No |  |
| Structured palliative care consultation and booklet “Advanced Dementia: A Guide for Families” | Care partners of individuals living with advanced dementia | Hanson et al. (2019) | Walsh et al. (2021)  Geddis-Regan et al. (2021) | Yes | <https://www.agingresources.com/wp-content/uploads/2018/08/DementiaGuide-final-2015.pdf> |
| Palliative consult intervention by physician and palliative care social worker | Care partners of individuals living with advanced dementia | Reinhardt et al. (2014) | Bryant et al. (2019)  Kelly et al. (2019) | No |  |
| Nurse-delivered adapted version of the “UK National Health Service Preferred Priorities of Care” | Care partners of individuals living with advanced dementia | Sampson et al. (2011) | Lord et al. (2015) | Yes | <https://www.stlukes-hospice.org.uk/wp-content/uploads/2017/06/accessible-version.pdf>  <https://www.choiceforum.org/docs/ppc.pdf> |
| Audiovisual booklet “Making Choices: long-term tube feeding placement in elderly patients” and worksheet  Three updates and three cultural adaptations, including the “International Patient Decision Aid Standards (IPDAS) statement” | Original: Care partners of individuals 65+ yrs  Update 1: Dyads of care partners and individuals living with advanced dementia  Updates 2 and 3: Care partners of individuals living with advanced dementia | Original: Mitchell et al. (2001)  Update 1: Hanson et al. (2010, 2011)  Update 2: Snyder et al. (2013)  Update 3: Ersek et al. (2014)  International IPDAS: Elwyn et al. (2006)  Portuguese adaptation: Derech and Neves (2021)  Japanese adaptation: Kuraoka and Nakayama (2014) | Cardona-Morrell et al. (2017)  Ho et al. (2021)  Davies et al. (2019)  Lord et al. (2015)  Pei et al. (2022)  Austin et al. (2015)  Kelly et al. (2019)  Petriwskyj et al. (2013)  Geddis-Regan et al. (2021)  Walsh et al. (2021)  Xie et al. (2018)  Backhaus et al. (2020) | Yes | <https://decisionaid.ohri.ca/docs/Tube_Feeding_DA/PDF/TubeFeeding.pdf>  Narration:  <https://decisionaid.ohri.ca/docs/Tube_Feeding_DA/PDF/TubeFeedingNarration.pdf> |
| “Guiding Options for Living with Dementia (GOLD)” | Care partners of individuals living with dementia | Stirling et al. (2012) | Austin et al. (2015)  Davies et al. (2019)  Ho et al. (2021)  Lord et al. (2015)  Mattos et al. (2023) | Yes | <https://figshare.utas.edu.au/ndownloader/files/40913426> |
| DECIDE manual: workbook and support of a decision coach | Care partners | Lord et al. (2017) | Davies et al. (2019)  Ho et al. (2021) | Yes | See appendix 28, page 248: <https://discovery.ucl.ac.uk/id/eprint/1522519/7/Lord_Kathryn_FINAL_KlordPhDThesis_Aug2016.pdf.%20REDACTED.pdf> |
| “At the Crossroads” | Care partners of individuals living with mild cognitive impairment, Alzheimer’s disease, or related dementia | Stern et al. (2008) | Davis and Ohman (2017)  Rapoport et al. (2012) | No |  |
| In-person session with a nurse or social worker, and manual | Care partners | Lingler et al. (2016) | Gench et al. (2021) | No |  |
| “ComputerLink” | Care partners | Bass et al. (1998)  Brenan et al. (1995) | Hopwood et al. (2018) | No |  |
| “AlzMed” booklet and website | Care partners | Zimmerman et al. (2018) | Gench et al. (2021) | Book: yes (for purchase)  Website: no | Book: <https://www.amazon.com/Alzheimers-Medical-Advisor-Caregivers-Experienced/dp/1934716669> |
| “Building Better Caregiver (BBC)” internet-based workshop | Care partners | Lorig et al. (2012) | Hopwood et al. (2018) | Yes | <https://va.buildingbettercaregivers.org/> |
| “European eHealthMonitor project Dementia Portal (eHM-DP)” | Care partners (informal and professional) | Schaller et al. (2015, 2016) | Hopwood et al. (2018) | No |  |
| “Palliative and Therapeutic Harmonization (PATH) program” | Individuals of 65+ yrs and their care partners | Moorhouse and Mallery (2012) | Kelly et al. (2019) | Yes | <https://pathclinic.ca/> |
| 2-minute video | Individuals living with dementia and their care partners | Volandes et al. (2008, 2009B, 2009B, 2011) | Davies et al. (2019)  Austin et al. (2015) | No |  |
| 18-minute video (2-min video updated) and ACP consultation | Individuals living with moderate-to-severe dementia and their care partners (50% African American) | Einterz et al. (2014) | Cardona-Morrell et al. (2017)  Davies et al. (2019)  Kelly et al. (2019)  Xie et al. (2018)  Ho et al. (2021) | No |  |
| 12-minute video | Individuals living with advanced dementia and their care partners | Mitchell et al. (2018) | Tunnard et al. (2022) | No |  |
| “Taking Control of Alzheimer’s Disease” | Individuals living with early dementia and their care partners | Silverstein and Sherman (2010)  Roberts and Silverio (2009) | Kelly et al. (2019) | No |  |
| “The Early Stage Memory Loss seminar” | Individuals living with dementia and their care partners | Logsdon et al. (2007) | Kelly et al. (2019) | No |  |
| Seminar about different types of ACP documentation | Individuals living with dementia and their care partners | Lewis et al. (2015) | Kelly et al. (2019) | No |  |
| “Let Me Decide” | Individuals living with dementia and their care partners | Molloy et al. (2000)  Caplan et al. (2006) | Kelly et al. (2019)  Robinson et al. (2012) | Yes (booked based on intervention) | <https://www.renaud-bray.com/Livres_Produit.aspx?id=722903&def=Let+me+decide%2CMOLLOY%2C+WILLIAM%2C0143055496> |
| “Preserving Identity and Planning for Advance Care (PIPAC)” | Individuals living with early dementia and their care partners | Hilgeman et al. (2014) | Kelly et al. (2019) | No |  |
| “Support, Health, Activities, Resources, and Education (SHARE)” | Individuals living with early dementia and their care partners | Whitlatch et al. (2019) | Geddis-Regan et al. (2021) | No |  |
| Discussion-based based intervention | Individuals living with dementia and their care partners | Meyer et al. (2019) | Mattos et al. (2023) | No |  |
| “Talking Mats” | Individuals living with dementia and their care partners | Murphy and Oliver (2013)  Dutch version: Reitz and Dalemans (2016) | Daly et al. (2018)  Davis et al. (2017)  Mattos et al. (2023) | Yes | <https://www.talkingmats.com/> |
| “DecideGuide” | Individuals living with dementia and their care partners (informal and professional) | Span et al. (2015)  Span (2016) | Hopwood et al. (2018)  Daly et al. (2018)  Mattos et al. (2023) | No |  |
| Five 6-to-10-minute videos | Individuals living with advanced dementia | Mitchell et al. (2020) | Tunnard et al. (2022) | No |  |
| “Driving with Dementia Decision Aid (DDDA)” | Individuals living with dementia | Carmody et al. (2014) | Ho et al. (2021) | Yes | <https://www.drivinganddementia.ca/uploads/DDDA_BOOKLET_CANADA_Jun%2019%202023.pdf> |
| Web-based intervention for decision-making based on personal values and preferences | 60+ yrs individuals living with mild cognitive impairment | Bogza et al. (2020) | Ho et al. (2021) | No |  |
